# An Optimised Monophasic Faecal Extraction Method for LC-MS Analysis and its Application in Gastrointestinal Disease

**DOI:** 10.1101/2022.09.19.22280109

**Authors:** Patricia E. Kelly, H Jene Ng, Gillian Farrell, Shona McKirdy, Richard K. Russell, Richard Hansen, Zahra Rattray, Konstantinos Gerasimidis, Nicholas J.W. Rattray

**Affiliations:** Strathclyde Institute of Pharmacy and Biomedical Sciences (SIPBS), University of Strathclyde, Glasgow, United Kingdom; School of Medicine, Dentistry & Nursing, University of Glasgow, Glasgow Royal Infirmary, United Kingdom, G31 2ER; Royal Hospital for Children and Young People, 50 Little France Crescent, Edinburgh EH16 4TJ; Royal Hospital for Children, 1345 Govan Road, Glasgow G52 4TF; Bacteria, Immunology, Nutrition, Gastroenterology and Omics Group, University of Glasgow; Glasgow, UK

## Abstract

Liquid chromatography coupled with mass spectrometry (LC-MS) metabolomic approaches are widely used to investigate underlying pathogenesis of gastrointestinal disease and mechanism of action of treatments. However, a standardised method for extracting metabolites from faecal samples for large-scale metabolomic studies is yet to be defined. Current methods often rely on biphasic extractions using harmful halogenated solvents, making automation and large-scale studies challenging. The present study reports an optimised monophasic faecal extraction protocol that is suitable for untargeted and targeted LC-MS analyses. The impact of several experimental parameters, including sample weight, extraction solvent, cellular disruption method, and sample-to-solvent ratio were investigated. It is suggested that a 50 mg freeze-dried faecal sample should be used in a methanol extraction (1:20) using bead beating as the means of cell disruption. This is revealed by a significant increase in number of metabolites detected, improved signal intensity, and wide metabolic coverage given by each of the above extraction parameters. Finally, we addressed the applicability of the method on faecal samples from patients with Crohn’s disease (CD) and coeliac disease (CoD), two distinct chronic gastrointestinal diseases involving metabolic perturbations. Untargeted and targeted metabolomic analysis demonstrated the ability of the developed method to detect and stratify metabolites extracted from patient groups and healthy controls (HC), highlighting characteristic changes in the faecal metabolome according to disease. The method developed is therefore suitable for the analysis of patients with gastrointestinal disease and can be used to detect and distinguish differences in the metabolomes of CD, CoD, and HC.

## 1. Introduction

Metabolomics is a powerful tool for detecting small molecule cellular and microbial products. Through the reflection of active physiological mechanisms, metabolite characterisation and quantification can give critical insights into human health and disease. The large abundance and diversity of metabolites that are present in human faecal samples, as given by the identification of over 6700 faecal metabolites on the Human Metabolome Database (HMBD - https://hmdb.ca/) [1], provides an ideal target for metabolomic analysis [2] and thus allows insights into outcomes of gut-microbial interactions and dietary impacts on disease [3]. Accumulating evidence indicating the involvement of the gut metabolome in a multitude of diseases [4-6] has propelled an intense interest in the role of faecal metabolites under certain environments. The accurate quantification of metabolites in faecal samples therefore holds value in a wide range of research areas. A clear role of faecal metabolomics has been demonstrated in the field of gastrointestinal disease, including inflammatory bowel disease (IBD) [7] and coeliac disease (CoD) [8]. Although the aetiology of such diseases remains elusive, shifts in metabolic profile are associated with disease activity and may represent central components of pathogenesis [9-12]. Irrespective, detection of altered patient metabolites may help unravel underlying disease mechanisms or reveal new diagnostic or prognostic markers of clinical utility.

Liquid-chromatography mass spectrometry (LC-MS) is a popular metabolite analysis technique due to its high sensitivity and selectivity. Sample preparation and pre-treatment is a vital stage of the LC-MS workflow, providing the scaffolding to support metabolite detection. The experimental framework therefore shapes the biological interpretation of a metabolomics study and so it is crucial to consider best practices regarding specific study aims. Certain challenges are inherent in the sample preparation phase, such as the large physio-chemical variation of the target metabolite pool, technical and environmental variation, and the complex and heterogeneous nature of human faeces. This brings difficulties in standardizing metabolomic methods, which is evident in the lack of “gold standard” metabolite extraction procedures. As the effective and reliable identification of metabolites is largely dependent on the extraction method used, it is imperative to consider sample preparation when comparing results between studies. To date, previous studies have addressed some of the challenges associated with metabolomic sample preparation [13-15], however these are mainly based on biphasic extraction protocols with limitations in scalability: While efficient biphasic extraction systems for faecal analysis contribute towards protocol standardisation, they are associated with complicated handling due to the requirement for phase separation. It can therefore be challenging to utilise two-phase protocols in large scale clinical studies, with further limitations in protocol automation. With the increasing demand for translating metabolomics data into meaningful clinical output, one major requirement for bridging the bench to bedside gap is the use of large population studies. It is therefore also important to optimise less-complex monophasic extraction protocols that can be used as an alternative to classical biphasic protocols for LC-MS analysis. Moreover, the applicability of metabolite extraction in the context of gastrointestinal disease requires further acknowledgement. Thus, the present study has the goal of advancing a method for monophasic metabolite extraction that can be easily implemented in large scale clinical studies investigating gastrointestinal disease. To the best of our knowledge, there is no current documentation on optimal extraction methods for IBD or CoD samples for LC-MS analysis. There is an important unmet requirement for the effect of faecal sample type to be explored, which is exemplified here in the comparison between gastrointestinal disease and the non-disease state.

Herein, we evaluate different faecal extraction methods for metabolomic measurements in human faecal samples from healthy individuals, Crohn’s Disease (CD) and CoD patients. A range of trial experiments were performed to determine the optimal sample weight, extraction solvent, disruption method, and sample-to-solvent ratio using LC-MS. The overall aim of this study is twofold; firstly, to optimise metabolite extraction parameters for faecal samples and secondly, to determine whether this optimised extraction protocol is suitable for analysis of samples from patients with gastrointestinal disease. To capture the large quantity of metabolites and ensure maximal coverage in the method development phase, untargeted metabolomic analysis was performed to assess each sample parameter. Targeted metabolomic analysis was subsequently applied to assess method suitability in patients with disease.

## 2. Materials and Methods

### 2.1 Overall Experimental Workflow

The overall experimental workflow including extraction, LC-MS metabolite measurement, data analysis, and data quantification and interpretation are shown Supplementary Information Figure S1).

### 2.2 Ethics Statement

All participants and their carers provided written informed consent. The study was approved by the NHS West of Scotland Research Ethics Committee (Ref: 11/WS/0006) for the study in patients with coeliac disease and the Yorkhill Research Ethics Committee (Reb: 05/S0708/66) for the study in patients with Crohn’s disease.

### 2.3 Faecal Samples

Faecal samples were collected for metabolomic analysis and freeze-dried prior to processing. Samples were stored at -80 **°**C until metabolite extraction and again until LCMS analysis. Samples were kept on ice during transportation.

### 2.4 Chemicals and Reagents

LC-MS grade methanol (MeOH), acetonitrile (ACN), chloroform (CHCl_3_), and water (H_2_O) were purchased from Fisher Scientific (Geel, Belgium). LC-MS grade formic acid was purchased from Thermo Scientific (Czech Republic).

### 2.5 Extraction Protocol

Freeze-dried faecal sample was added to extraction solvent and cells were disrupted using bead beating (FastPrep 24 MP Biomedicals), sonication, and freeze-thaw lysis methods. Samples were then centrifuged at 13,000 g for 15 minutes and the supernatant recovered. Samples were lyophilised for 36 hours and stored at -80 **°**C until further processing. Reconstitution was performed in 250 μL 50/50 H_2_O: acetonitrile (ACN), vortexed for 1 minute and centrifuged at 15,000 g for 15 minutes, and aliquots transferred into glass vials for MS analysis. Quality control (QC) samples were prepared by pooling samples across all groups undergoing simultaneous analysis. Solvent blanks and QC samples were entered at the beginning of every analytical run and after every five samples in each batch over the course of the study to assess background in the system and detect potential contaminations.

**Table 1.**
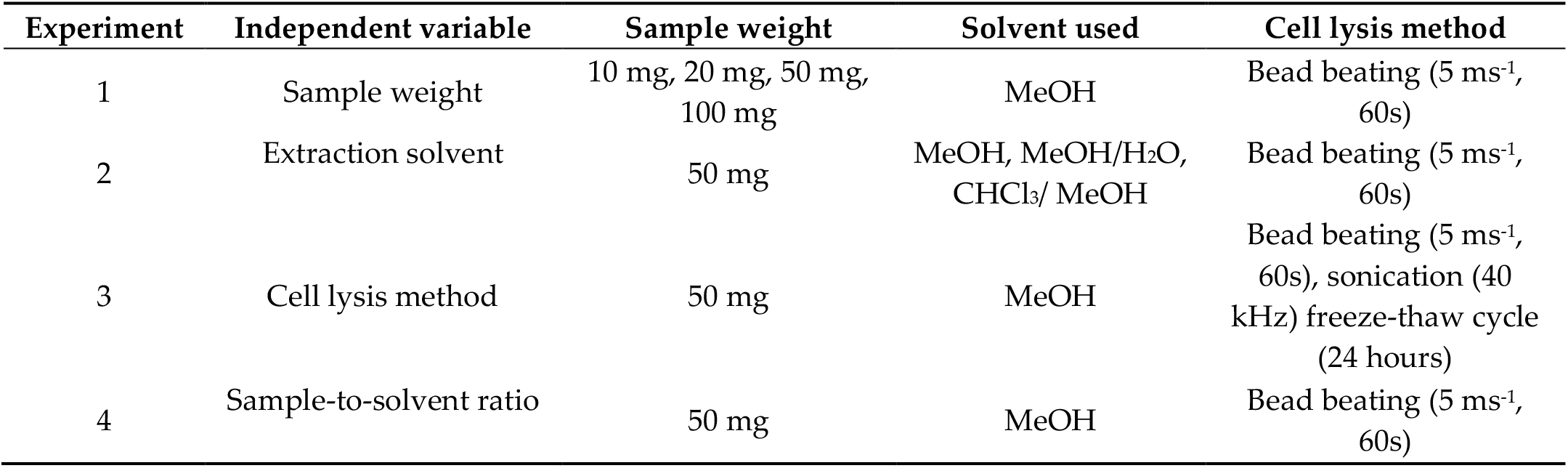
Details of experimental conditions for each extraction parameter.

| Experiment | Independent variable | Sample weight | Solvent used | Cell lysis method |
| --- | --- | --- | --- | --- |
| 1 | Sample weight | 10 mg, 20 mg, 50 mg, 100 mg | MeOH | Bead beating (5 ms<sup>-1</sup>, 60s) |
| 2 | Extraction solvent | 50 mg | MeOH, MeOH/H<sub>2</sub>O, CHCl<sub>3</sub>/ MeOH | Bead beating (5 ms<sup>-1</sup>, 60s) |
| 3 | Cell lysis method | 50 mg | MeOH | Bead beating (5 ms<sup>-1</sup>, 60s), sonication (40 kHz) freeze-thaw cycle (24 hours) |
| 4 | Sample-to-solvent ratio | 50 mg | MeOH | Bead beating (5 ms<sup>-1</sup>, 60s) |

### 2.6 Untargeted LC-MS Metabolite Measurement

Untargeted metabolomic analysis was performed on an ultra-high performance liquid chromatography UHPLC) system (ThermoFisher Scientific) coupled to an Orbitrap Exploris 240 (ThermoFisher Scientific) mass spectrometer. Chromatographic separation was performed on a Vanquish Accucore C18 + UHPLC analytical column (ThermoScientific, 100 mm x 2.1 mm, 2.6 μM) at a flow rate of 400 μL min ^-1^. Mobile phase A was composed of 99.9% water + 0.1% formic acid and mobile B was composed of 99.99% MeOH + 0.1% formic acid. Electrospray ionisation (ESI) was used as the ionisation method, set at 3900 V and 2500 V for positive and negative mode, respectively. The elution gradient used can be found in Supplementary Information Table S1. The source-dependent parameters were operated under the following conditions: sheath gas, 40 Arb; auxiliary gas, 10 Arb; sweep gas, 1 Arb; ion transfer tube temperature, 300 **°**C; vaporiser temperature, 280 **°**C. Instrument calibration was performed using Pierce™ FlexMix™ calibration solution (Thermo Scientific) and ran under vendor recommended settings. MS data collection was performed in a top-5 data dependent acquisition mode (DDA).

### 2.7 Targeted LC-MS Metabolite Measurement

Targeted metabolomic analysis was performed on an ultra-high-performance liquid chromatography (UHPLC) system coupled to a triple quadrupole mass spectrometer (Shimadzu 8060NX). The method used for metabolite detection and quantification was provided by the vendor: Primary Metabolites LC/MS/MS Method Package version 2.0 The method is designed to detect 97 metabolites. The list of 97 detected metabolites is shown in Supplementary Information Table S3. Chromatographic separation was performed on a pentafluorophenylpropyl (PFPP) + UHPLC analytical column (Merck, 150 mm x 2.1 mm, 3 μM) at a flow rate of 400 μL min ^-1^. Mobile phase A was composed of 99.9% water + 0.1% formic acid and mobile B was composed of 99.9% acetonitrile + 0.1% formic acid. Electrospray ionisation (ESI) was used as the ionisation method, set at 3900 V and 2500 V for positive and negative mode, respectively. The source-dependent parameters were operated under the following conditions: column oven temperature, 40 **°**C; nebulising gas flow rate, 3.0 L min^-1^, drying gas flow rate, 10 L min^-1^, desolvation line temperature, 250 **°**C; block heater temperature, 400 **°**C. The elution gradient used can be found in Supplementary Information Table S2.

### 2.8 Method Application

We applied the method to three biological groups: CD patients, CoD patients, and HCs. CD patients were undergoing varying forms of treatment and CoD patients were following a gluten-free diet. HCs were defined as individuals with the absence of gastrointestinal disease. Untargeted and targeted metabolomic analyses were applied to the sample sets combined after randomisation.

**Table 2.**
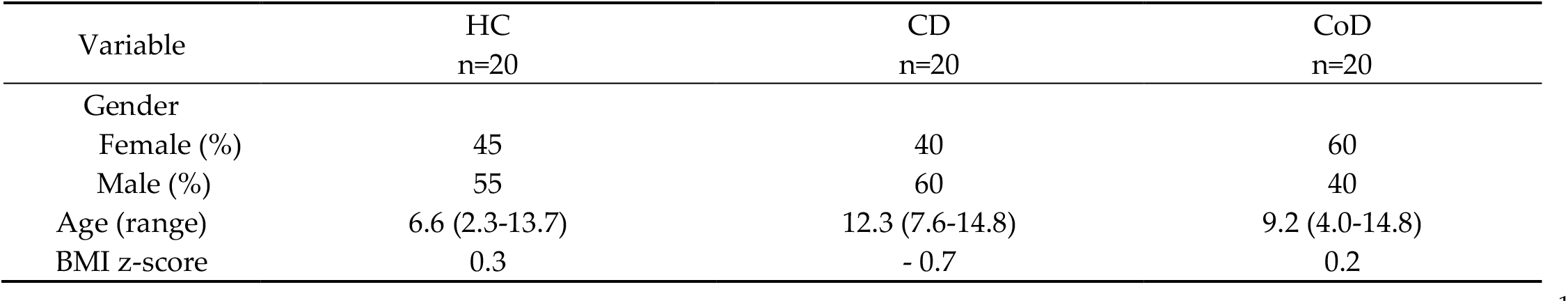
Table of patient demographics. HC, healthy control; CD, Crohn’s disease, CoD, Coeliac disease.

### 2.9 Mass Spectrometry Data Processing

For the processing of untargeted metabolomics data, Thermo Scientific Xcalibur format raw data files (.RAW) were imported into Compound Discoverer software (version 3.2). Details of the workflow for analysis in Compound Discoverer is included in Supplementary Information Table S4. Targeted metabolomics data were converted from Shimazdu vendor format (.lcd) to mzML format. A data matrix of identified metabolites and associated peak areas was constructed and processed using R-Studio v 3.5.2.

### 2.10 Data and Statistical Analysis

For untargeted analysis, multivariate statistical analysis was performed using Compound Discoverer software 3.2. For targeted analysis, multivariate statistical analyses were performed using Lab Profiling Solutions software version 1.1 and R-Studio. Principal component analysis (PCA) was used to reduce data dimensionality, through which PCs were calculated using prcomp function and PCA scores plots were generated using the following packages in R: ggplot2, ggfortify, grid, and gridExtra. Differential analysis using volcano plots allowed significant differences between groups to be determined. Univariate statistical analyses were performed using unpaired t-test and one-way ANOVA, with the level of significance set at p < 0.05. Central network analysis was performed in R-studio using the igraph package.

### 2.11 Metabolite Identification

Inclusion criteria for metabolite identification were set and applied to refine the total number of features (Supplementary Information Figure S2). The resulting list contained only metabolites within a 10 ppm mass accuracy range and with a full predicted composition and ChemSpider match. Contaminations were excluded by analysing 50:50 H_2_O/MeOH blank samples throughout the MS run. Metabolite annotation was performed manually and using the HMDB.

## 3. Results

For method development, metabolites were measured in freeze-dried faecal samples obtained from healthy participants. The metabolic output was first measured by PCA to observe any differences in the overall metabolic signature obtained from each method. Further statistical analysis was performed for data quantification by calculating the number of m/z features, identified metabolites, signal intensity, and metabolic coverage.

### 3.1. Analysis of Sample Weight

A significantly higher number of m/z features were detected through the positive ionisation acquisition mode in comparison to the negative ionisation mode (Supplementary Information Figure S3). For this reason, the positive ionisation mode was subsequently used for further analysis of experimental parameters. Metabolite extraction from 10 mg and 100 mg samples were unsuitable for metabolomic analysis and therefore not included in the results. Metabolites were successfully extracted from 20 mg and 50 mg samples and measured using LC-MS. Multivariate statistical analysis demonstrated clear separation of the two sample weight groups (**Fig. 1**). 50 mg samples showed a significantly higher mean number of m/z features and mean number of identified metabolites in comparison to 20 mg samples. Furthermore, the mean signal intensity given by 50 mg samples (2.5×10^7^) was significantly increased compared to 20 mg samples (1.1×10^7^). Levels of altered metabolites were calculated from the differential analysis of sample weight and expressed as the percentage of significantly increased metabolites between groups. It was demonstrated that 59.6% of identified metabolites were found at significantly increased levels in 50 mg samples compared to 20 mg samples (Supplementary Information Figure S4). All metabolite classes were detected from both sample groups, with similar compositions of chemical class observed. A comparison of the total number of metabolites per chemical class identified using each sample weight is shown in Supplementary Information Figure S5.

**Figure 1.**
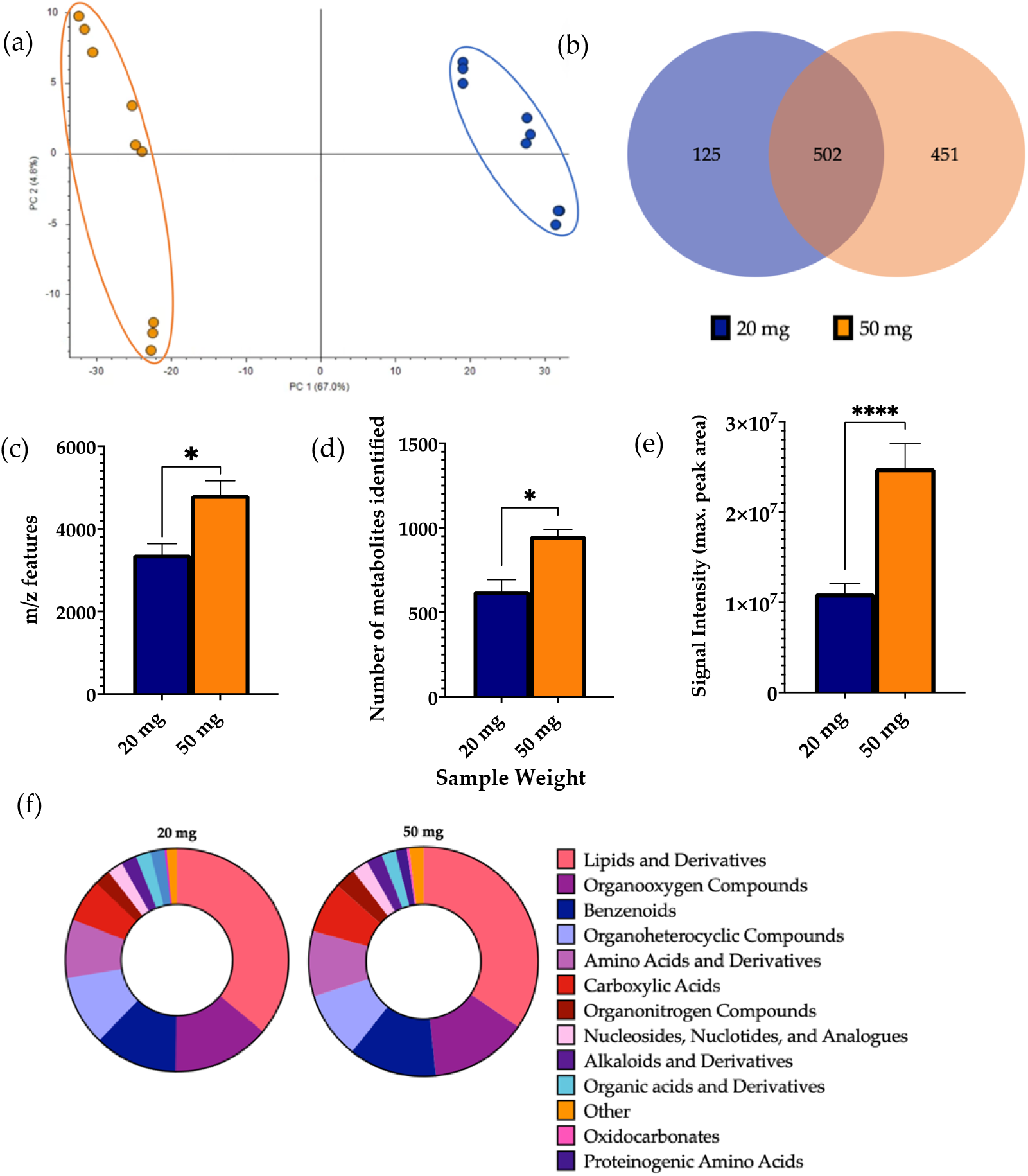
Effect of sample weight on features of metabolomic analysis. 1 μL of 20 mg and 50 mg sample was injected onto a C18 column (n=3), performed in triplicate. (a) Multivariate analysis of metabolomic profiles obtained as a function of sample weight. PCA score plots demonstrating extracted faecal metabolites between different sample weights. Discrimination between 20 mg (blue) and 50 mg (orange) samples was characterised by a variability of 71.6%. (b) Venn diagram of the mean number of identified metabolites between each method. (c) The total number of m/z features and (d) total number of identified metabolites were calculated in positive ionisation mode and (e) the overall mean signal intensity of each sample weight was assessed. (f) Metabolite class quantification demonstrating the faecal metabolome patterns identified by chemical class in 20 mg and 50 mg samples. Bar chart data are expressed as mean ± SEM and statistical significance was assessed using an unpaired t-test. *p < 0.05, **** p < 0.0001.

### 3.2 Analysis of Extraction Solvent

Multivariate statistical analysis demonstrated clear separation of the extraction solvents (**Fig. 2**). Using 100% MeOH gave a significantly higher number of m/z features in comparison to MeOH/H_2_O and a significantly higher number of identified metabolites than both MeOH/H2O and CHCl_3_/ MeOH. No significant differences were observed in the signal intensity between the extraction solvents. Differential analysis revealed a significant increase in the levels of 28.5% and 9.3% of identified metabolites using MeOH as the extraction solvent in comparison to MeOH/H_2_O and CHCl_3_/ MeOH, respectively (Supplementary Information Figure S6). 25.8% of identified metabolites were found at significantly increased levels in CHCl_3_/ MeOH compared to MeOH/H_2_O. MeOH extractions additionally had a significantly increased number of lipids compared to MeOH/H_2_O extractions. Again, all metabolite classes were detected from all extraction solvents, with a similar structure of metabolite classification. A comparison of the total number of metabolites per chemical class identified using each extraction solvent is shown in Supplementary Information Figure S7.

**Figure 2.**
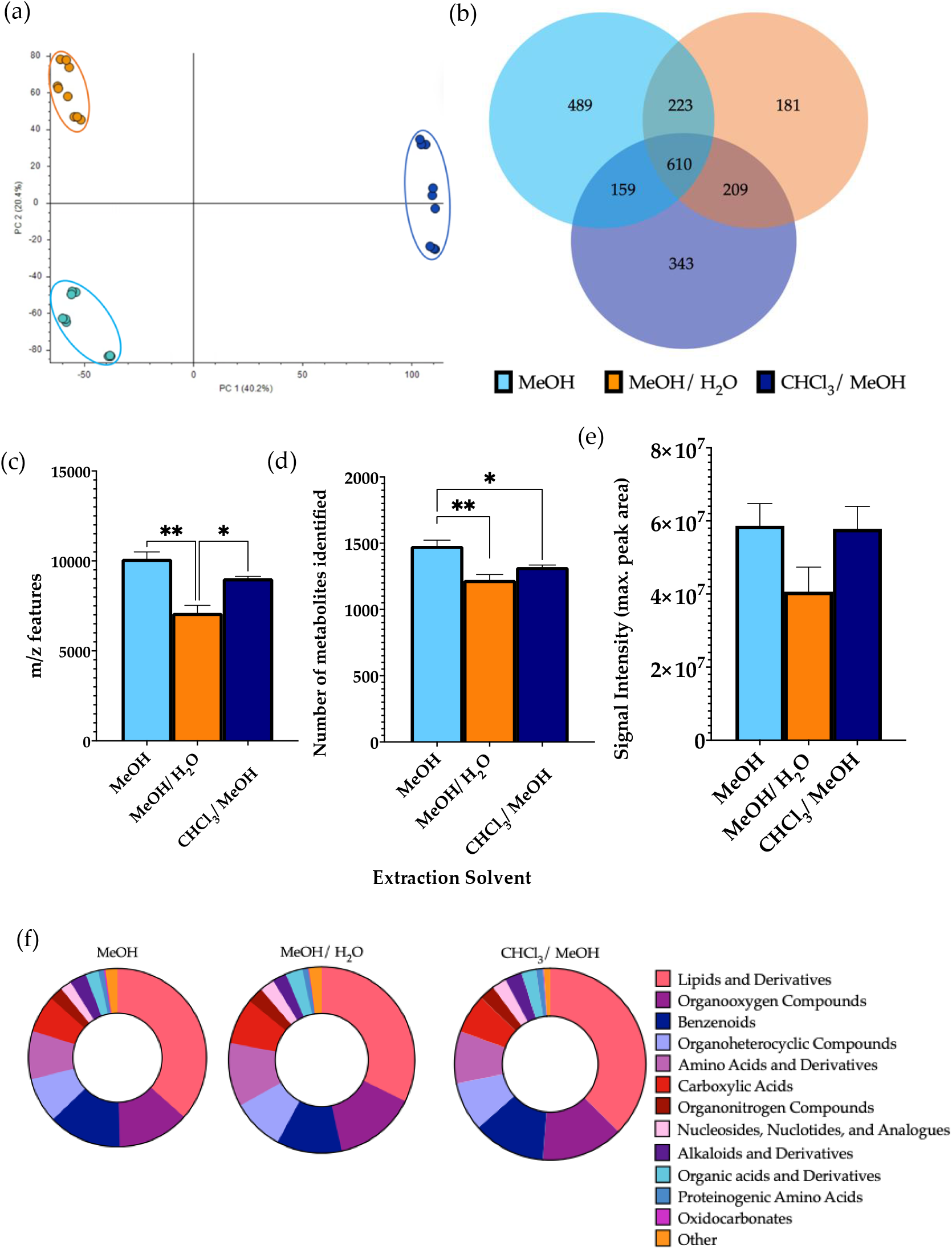
Effect of extraction solvents, MeOH, MeOH/H_2_O, and CHCl_3_/ MeOH, on features of metabolomic analysis. 1 μL of each extraction sample was injected onto a C18 column (n=3), performed in triplicate. (a) Multivariate analysis of metabolomic profiles obtained as a function of extraction solvent. PCA score plots demonstrating extracted faecal metabolites between different extraction solvents. Discrimination between extraction solvents MeOH (light blue), MeOH/H_2_O (orange), and CHCl_3_/ MeOH (dark blue) was characterised by a variability of 40.2%. (b) Venn diagram of the mean number of identified metabolites between each method. (c) The total number of m/z features and total number of identified metabolites were calculated in positive ionisation mode and (e) the overall mean signal intensity of each extraction solvent was assessed. (f) Metabolite class quantification demonstrating the faecal metabolome patterns identified by chemical class in each extraction sample. Bar chart data were expressed as mean ± SEM and statistical significance was assessed using one-way ANOVA. *p < 0.05, ** p < 0.01.

### 3.3. Analysis of the Cellular Disruption Method

The choice of cellular disruption method affected the overall metabolic output, as shown by multivariate statistical analysis which demonstrated clear separation between the three groups (**Fig.3**). Bead beating extracted a significantly higher mean number of m/z features in comparison to freeze-thawing and a significantly higher number of identified metabolites than both sonication and freeze-thawing. No significant differences were observed in the signal intensity between lysis methods. A significant increase in the levels of 32.9% and 48.6% of identified metabolites were found using bead beating as the method of cellular disruption compared to sonication and freeze-thawing, respectively (Supplementary Information Figure S8). Of the metabolites identified, 26.2% were found at significantly increased levels in sonicated samples in comparison to freeze-thawing. Each disruption method allowed measurement of metabolites from all classification groups. While similar patterns of metabolite classification are shown between methods, it was shown that bead beating led to detection of a significantly increased number of lipids compared to the other lysis techniques. A comparison of the total number of metabolites per chemical class identified using each cellular disruption method is shown in Supplementary Information Figure S9.

**Figure 3.**
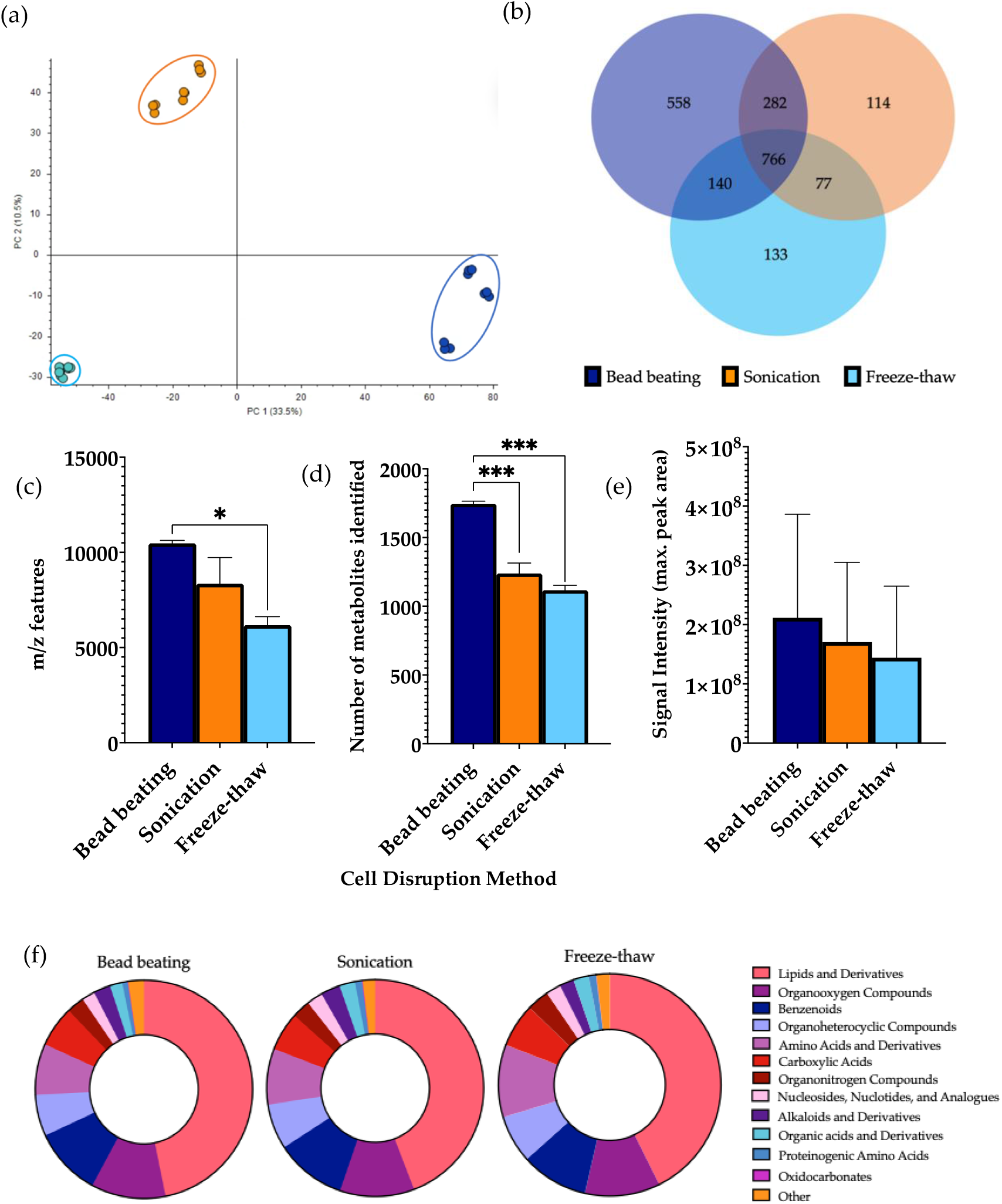
Effect of cellular disruption methods, bead beating, sonication, and freeze-thaw cycles, on features of metabolomic analysis. 1 μL of each extraction sample was injected onto a C18 column (n=3), performed in triplicate. (a) Multivariate analysis of metabolomic profiles obtained as a function of disruption method. PCA score plots demonstrating extracted faecal metabolites between bead beating, sonication, and freeze-thaw cycles. Discrimination between extraction solvents A, bead beating (dark blue); B, sonication (orange) and C, freeze-thaw cycles (light blue) was characterised by a variability of 33.5%. (b) Venn diagram of the mean number of identified metabolites between each method. (c) The total number of m/z features and (d) total number of identified metabolites were calculated in positive ionisation mode and (e) the overall mean signal intensity of each disruption method was assessed. (f) Metabolite class quantification demonstrating the faecal metabolome patterns identified by chemical class in each extraction sample. Bar chart data were expressed as mean ± SEM and statistical significance was assessed using a one-way ANOVA. *p < 0.05, **** p < 0.0001.

### 3.4. Analysis of Sample-to Solvent Ratio

Clear separation was observed between the three different sample-solvent ratios by PCA (**Fig.4**). Performing extractions using a ratio of 1:20 gave a significantly higher mean number of m/z features and identified metabolites than ratios of 1:5 and 1:10. Furthermore, a significant increase in the signal intensity of samples of a 1:20 ratio was observed in comparison to the other groups. A significant increase in the levels of 70.2% and 67.3% of identified metabolites were found using a ratio of 1:20 in comparison to ratios of 1:5 and 1:10, respectively (Supplementary Information Figure S10). 70.2% of identified metabolites were found at significantly increased levels in samples extracted using a ratio of 1:10 compared to 1:5. Amino acids and derivatives, lipids and derivates, organooxygen, and organoheterocyclic compounds were significantly increased in extractions carried out using a ratio of 1:20 compared to the other groups. Additionally, the overall composition according to chemical class of each sample remained similar between each group. A comparison of the total number of metabolites per chemical class identified using each sample-to-solvent ratio is shown in Supplementary Information Figure S11.

**Figure 4.**
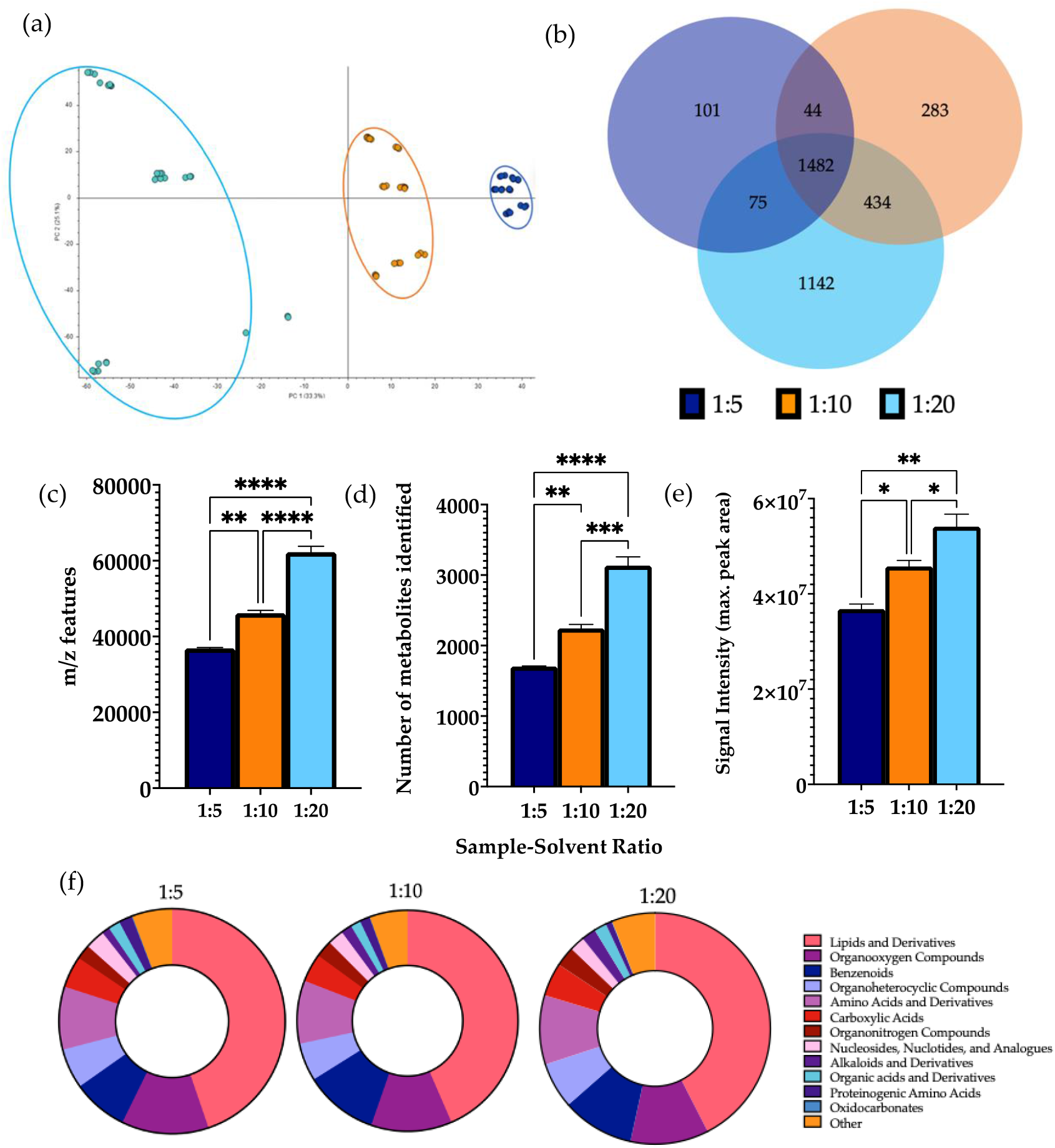
Effect of sample-solvent ratio on features of metabolomic analysis. 1 μL of each extraction sample was injected onto a C18 column (n=3), performed in triplicate. (a) Multivariate analysis of metabolomic profiles obtained as a function of sample-to-solvent ratio. PCA score plots demonstrating extracted faecal metabolites between different ratios. Discrimination between extraction solvents 1:5 (dark blue), 1:10 (orange) and 1:20 (light blue) was characterised by a variability of 33.3%. (b) Venn diagram of the mean number of identified metabolites between each method. (c) The total number of m/z features and (d) total number of identified metabolites were calculated in positive ionisation mode and (e) the overall mean signal intensity of each sample-to-solvent-ratio was assessed. (f) Metabolite class quantification demonstrating the faecal metabolome patterns identified by chemical class in each extraction sample. Bar chart data are expressed as mean ± SEM and statistical significance was assessed using a one-way ANOVA. **p < 0.01, *** p < 0.001, **** p < 0.0001.

Through exploration of the overall metabolite extraction efficiency through the optimisation process, it was observed that both the number of m/z features and identified metabolites significantly increased throughout stages of method optimisation with the improvement of each individual extraction parameter (Supplementary Information Figure S12).

### 3.5. Applicability of the Method to Patients with Gastrointestinal Disease

To assess the applicability of the developed method, we applied the protocol to CD, CoD, and HC groups and compared the metabolic differences. In an untargeted analysis, multivariate statistics demonstrated clear separation between CD samples and the other groups (**Fig.5**). A significant decrease in the levels of 72.3% of identified metabolites were found in CD samples compared to HCs, and 74.1% compared to CoD samples (Supplementary Information Figure S13). Of the metabolites identified, 27.1% were found at significantly decreased levels in CoD samples in comparison to HCs. Furthermore, targeted metabolomics analysis further confirmed the ability of the method to both detect and stratify metabolites extracted from faecal sample from patients with CD and CoD and healthy individuals. Multivariate analysis showed characteristic changes in the faecal metabolome between each of the groups (**Fig.6**).

**Figure 5.**
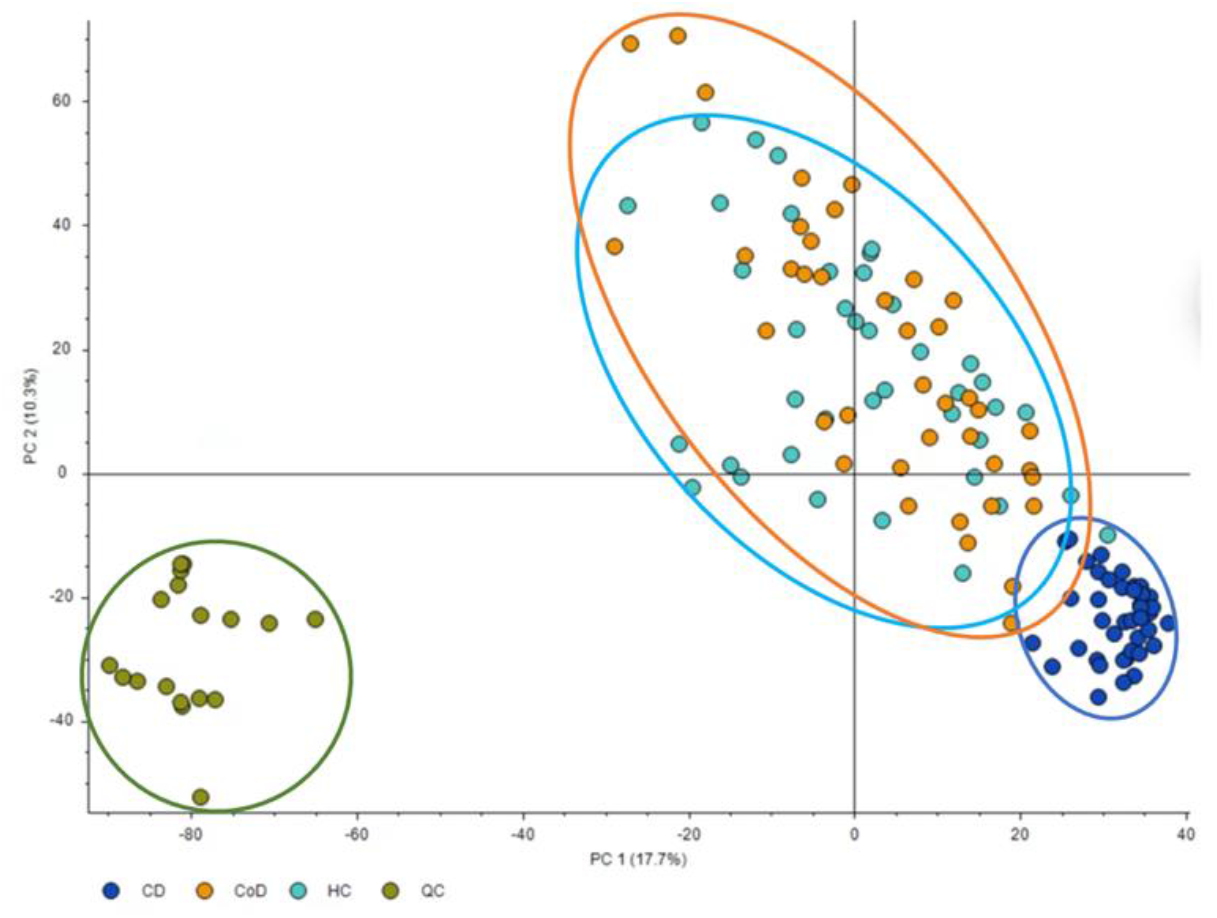
Multivariate analysis of metabolomic profiles based on untargeted analysis of gastrointestinal disease. PCA score plots demonstrating extracted faecal metabolites between patient groups. Principle Component 1 directionality describes the variance between CD (dark blue), CoD (orange) and HC (light blue) and explains 17.7% of the total variance of the data. QCs are shown in green. Samples are represented by individual datapoints, performed in triplicate (n=20).

**Figure 6.**
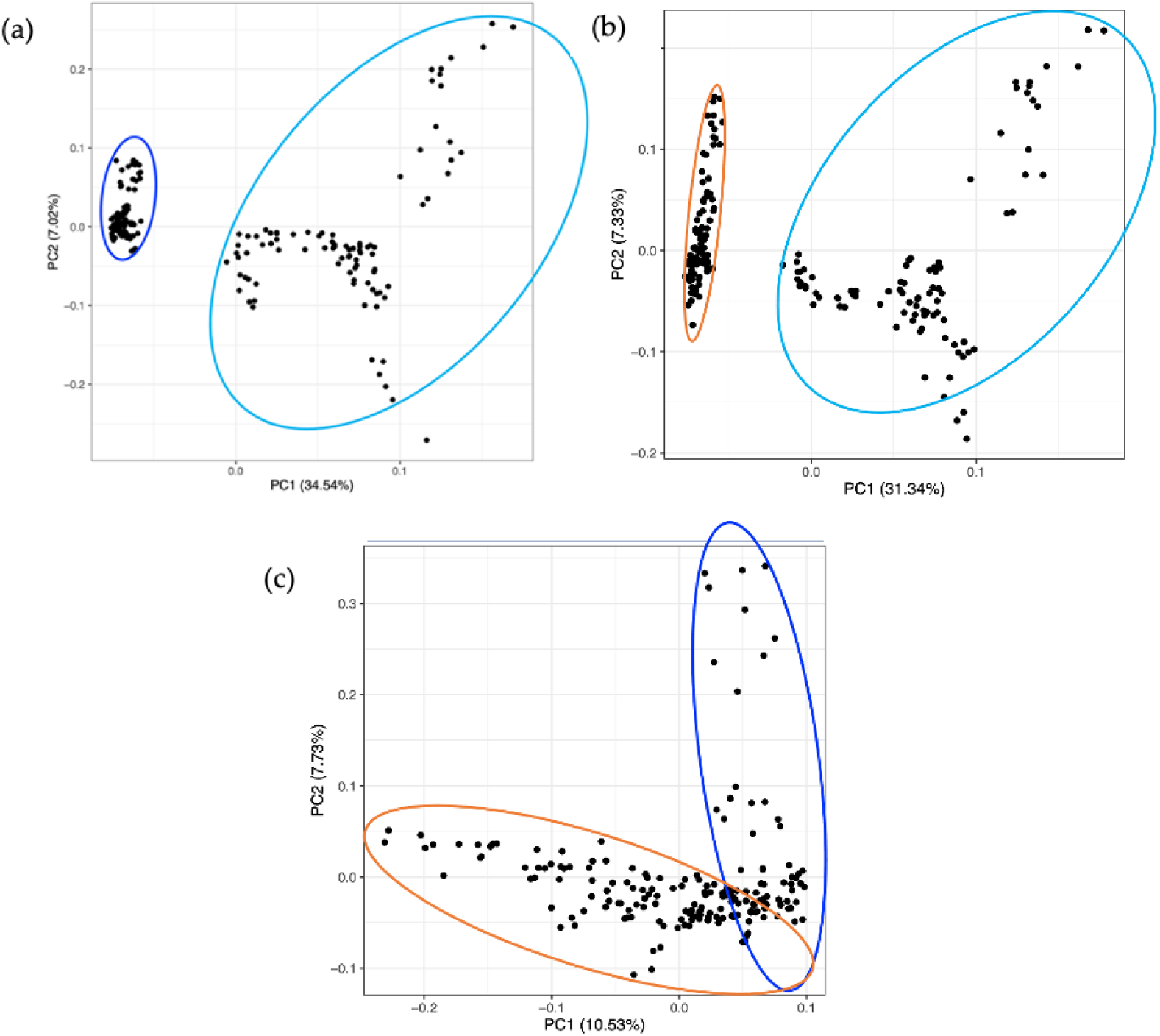
Multivariate analysis of the metabolomic profiles based on targeted analysis of gastrointestinal disease. PCA score plots demonstrating extracted faecal metabolites between CD (dark blue), CoD (orange) and HC (light blue). Discrimination between (a) CD vs. HC, (b) CoD vs. HC, and (c) CD vs. Co was characterised by variabilities of 34.5%, 31.3%, and 10.5%, respectively. Samples are represented by individual datapoints, performed in triplicate (n=20).

To ensure the present method was effective in the specific context of gastrointestinal disease, we carried out further analysis investigating metabolites that are important in IBD. Metabolites previously implicated in IBD pathology throughout the literature were identified, through a significant alteration of their levels in individuals with disease in comparison to healthy controls. A cross-study analysis was then carried out to assess whether the method developed in the present study was able to successfully detect those metabolites identified with involvement in IBD. It was found that the present method detected 75% of the metabolites identified in the IBD target list (**Fig.7**).

**Figure 7.**
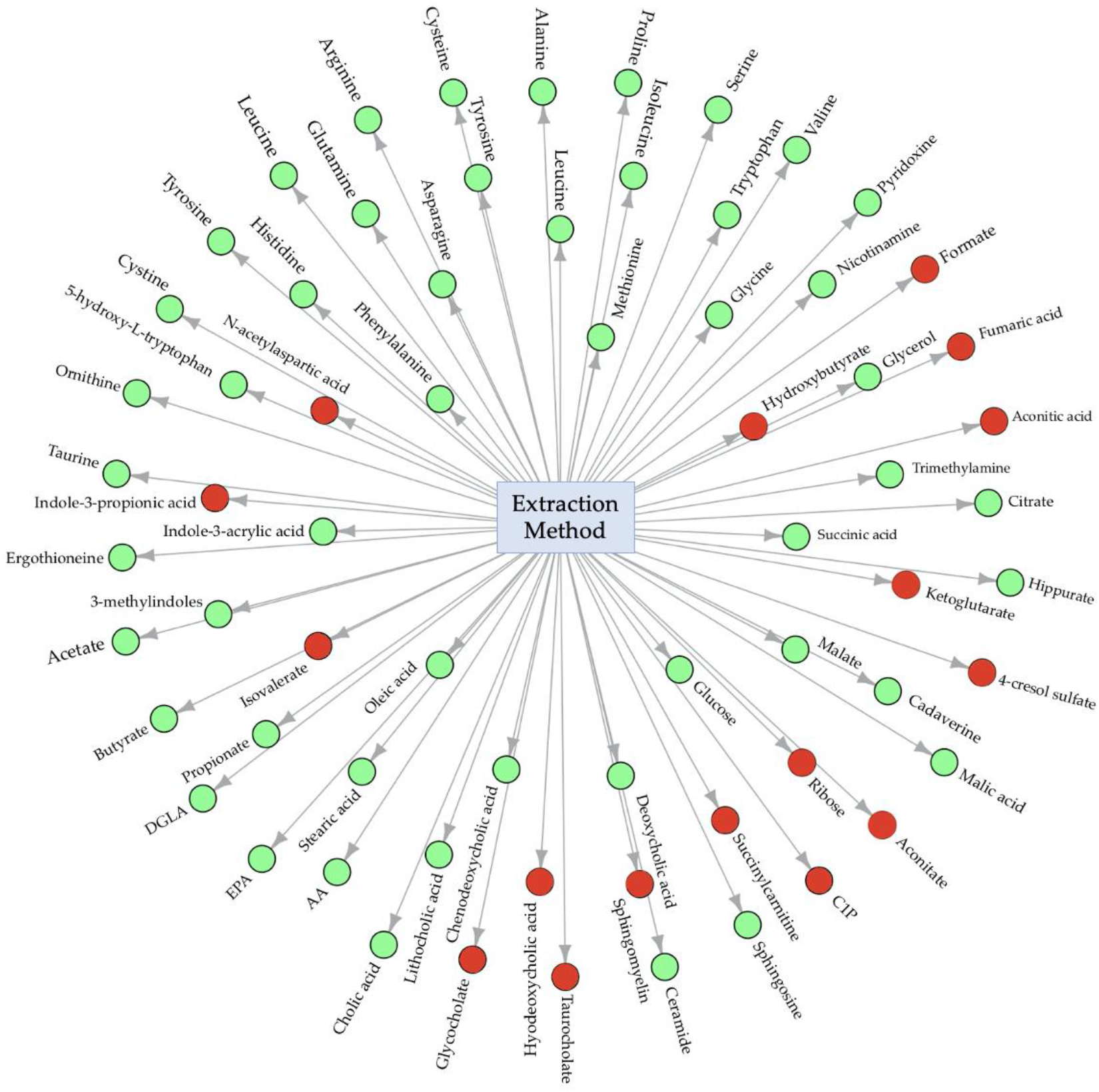
Central network analysis of developed metabolite extraction method. Circles shown in green represent metabolites successfully extracted using the developed method and circles shown in red represent metabolites not found using the developed method. C1P, Ceramide-1-phosphate; AA, Arachidonic acid; EPA, Eicosapentanenoic acid; DGLA, Dihomo-gamma linolenic acid.

## 4. Discussion

Since extraction methodology directly affects metabolite constitution within MS metabolomics experiments, it was important to optimise a range of experimental parameters and document the chemical coverage in faecal samples. To this end, the present study first aimed to assess parameters of maximal metabolic LC-MS output, utilising an untargeted metabolomics approach to allow fingerprinting of the total metabolite profile in samples. The ideal extraction protocol was therefore one that elucidated the greatest number of metabolites whilst minimizing interferences. As such, methods were evaluated by measuring the total number of metabolites detected using each protocol. Since we cannot assume that the number of features is equal to the number of correctly identified metabolites, due to unmatched features, blanks, and duplicate readings, further refinement methods were applied to allow for a more accurate evaluation of the protocols. Additionally, the markedly different characteristics of metabolites in the faecal metabolite pool brings challenges in extracting all the metabolites present in each sample. For this reason, it was important to assess the number of metabolites belonging to different metabolic classes from each method to ensure maximum chemical coverage. Feature annotation was performed to quantify and compare metabolite classifications between the extraction methods. As complete characterisation of the metabolome is not possible, a compromise will always exist in practice, however the multi-parameter method used in the present study allows method selection of the greatest coverage largely without sacrificing data quality.

Herein, we describe an optimised protocol for extraction of metabolites from human faecal samples, thus providing an efficient setup for subsequent metabolomic analysis. The method is recapitulated in the following stages: (1) 50 mg sample weighed out; (2) 1000 μL MeOH added to sample and cell lysed by bead beating; (3) samples lyophilised and stored at -80 **°**C until further processing; (4) reconstitution carried out in 50/50 ACN: H_2_O; (5) LC-MS analysis using 1 μL injection volume (**Fig.8**).

**Figure 8.**
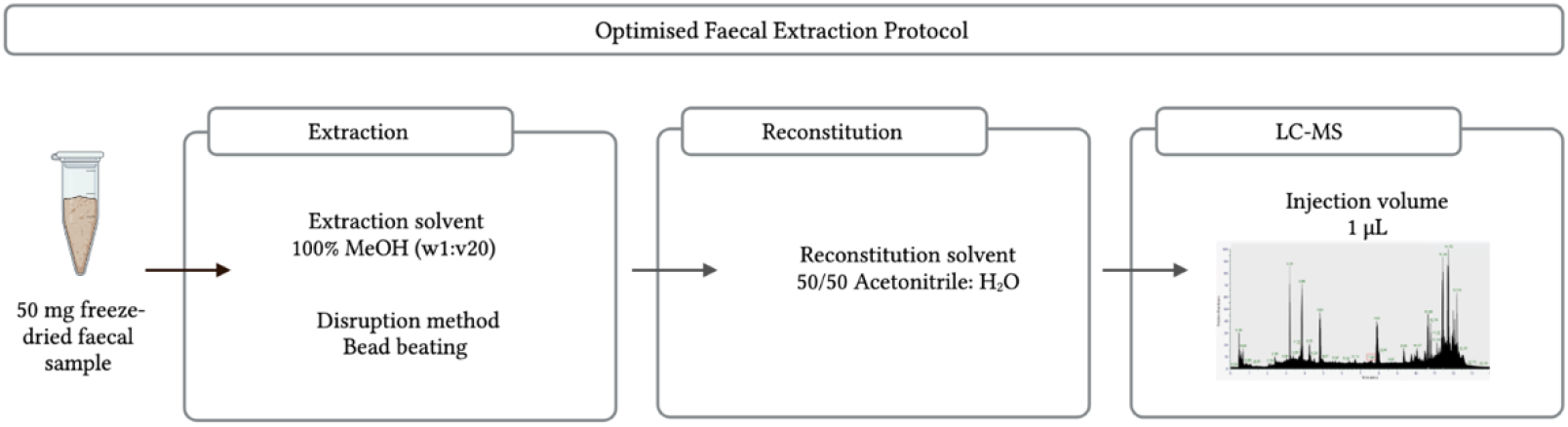
Summary of the developed methodology pipeline. Multi-parameter analysis showed that 50 mg samples give the strongest MS output, and from the extraction solvents analysed, MeOH is the most effective. Additionally, cellular metabolite release is optimal using bead beating as the cell lysis method. Combining optimised parameters provides an experimental protocol for faecal metabolite extraction that can be used for metabolomic analysis.

While examining the effect of sample weight, 10 mg samples were disregarded during the extraction process as the aliquots had very little extractable supernatant for subsequent processing. This was likely due to the sample being absorbed by the zirconium beads as the sample weight was too small for the solvent volume. During the reconstitution step, the 100 mg sample was also disregarded as there was too much particulate left undissolved. This is important as when run on the MS, sample particulate may crash the column and lead to instrument damage. The faeces weight-to-solvent ratio (100 μL of solvent for every 10 mg of sample) was therefore not sufficient for samples out with a 20-50 mg range. For this reason, we explored the impact of sample-solvent ratio on metabolic output in a further analysis (experiment 4). In consideration to this, for the assessment of sample weight, 20 mg and 50 mg samples were successfully extracted and metabolomic analysis was continued.

A clear separation was shown by the PCA comparing 20 mg and 50 mg samples, indicating the different metabolite profiles given by the two groups. Further analysis showed that 50 mg samples additionally contained an increased number of m/z features, identified metabolites, and signal intensity – this result was to be expected due to the increased levels of biomass in the 50 mg samples. It was also important to investigate whether the observed differences in metabolite numbers were reflected in the overall metabolic coverage. Thus, the identified metabolites were grouped according to their chemical classification, and calculation of the number of metabolites in each class was used as a measurement of metabolic coverage. This is essential for untargeted metabolomics experiments, as the analytical conditions should aim to detect a broad range of metabolites of different chemical properties that may be implicated in disease. As such, expansion of metabolic coverage is important to maximise information for hypothesis generation. From the classification analysis, it was revealed that metabolite class is conserved across sample weight. Using 50 mg faecal samples for metabolite extraction aligns with previously reported studies [3, 16-18], in which 50 mg samples were also used as the starting point for sample preparation and subsequent analysis. Based on findings of increased metabolite numbers without compromising metabolic coverage or signal intensity, it is reasonable to suggest that 50 mg samples are optimal for use in faecal extraction protocols.

Analysis of extraction solvent also showed a clear separation between protocols using MeOH, MeOH, MeOH/H_2_O, and CHCl_3_/ MeOH, with an increased number of m/z features and identified metabolites given by pure MeOH extractions. While it was shown that the number of lipids and derivatives were increased in the samples extracted using MeOH in comparison to the other groups, the overall metabolic coverage was very similar for all extraction solvents investigated. As maximal chemical coverage is largely maintained, it can again be noted that metabolite class is conserved across extraction solvents. As the use of pure MeOH increases overall metabolic features obtained from molecules across a wide range of different chemical properties, its use can therefore be recommended as the optimal solvent for faecal extraction. This result is in agreement with a recently reported study, where MeOH was chosen as the optimal solvent for extraction of metabolites from human faecal samples to assess gut health [19]. Furthermore, MeOH has been found to be the optimal extraction solvent in a range of metabolomics studies including investigation of dietary influences in faecal samples [3], serum metabolite profiling [20], and *Blastocystis*’ metabolism [21]. In comparison to one of the current most used extraction solvents, phosphate buffer saline (PBS) [22], the recognition of MeOH as an efficient organic buffer and resultant choice in a range of sample preparation methods may be attributed to effective protein denaturation [23] and multi-polarity chemical capture [24].

Cell lysis is the process of breaking down the cell membrane to release contents contained inside the cell for molecular analysis. Bead beating, sonication, and cycles of freeze-thawing are common techniques used to disrupt the cell, and a sense of uncertainty resides about optimal methodological choice. Samples that underwent cell lysis using bead beating contained a significantly higher number of m/z features than freeze-thawing and a significantly increased number of identified metabolites than both those with sonication and freeze-thawing. Moreover, cell disruption by bead beating had a significantly increased number of lipids compared to both other methods. Overall, these findings indicate that bead beating was the most effective cell lysis method for extracting metabolites from human faecal samples. Additional studies have found analogous findings, for example, one study showed that bead beating was the best method for cell disruption and subsequent extraction of both polar and non-polar compounds from platelet samples, as given by optimal extraction efficiencies [25]. Bead beating has also previously been used as the cell lysis method of choice in sample preparation of human feacal samples [26], as well as for gastrointestinal stromal tumour [27] and characterisation of tissue samples [28].

Sample-to-solvent ratio, as aforementioned, is vital not only to maximise the data obtained but also to ensure sufficient sample quality for LC-MS analysis so as not to cause blockage and instrument breakdown. This is particularly important for complex biomatrices such as faeces, which are composed of an abundance of organic and cellular material. The sample-to-solvent ratio therefore must allow extraction of large metabolite numbers that are compatible with LC-MS systems. Therefore, the metabolic output resulting from sample-to-solvent ratios of 1:5, 1:10, and 1:20 were assessed. Different metabolite quantification analyses identified a higher number of m/z features, identified metabolites, and signal intensity were given by samples using a sample-solvent ratio of 1:20 compared to the other tested ratios. Over 3000 m/z features were detected and putatively identified using the optimal procedure with a 1:20 sample-solvent ratio, which holds great promise for maximising capture of biological information in future metabolomic studies. It is important to note that this work is part of an ongoing effort to document the metabolites putatively identified in faecal samples which will in future will be built upon by the creation of a standards library and the additional use of pure standards. This will help work towards reporting standards of chemical analysis [29] and increase the confidence of identification.

Finally, we demonstrated the applicability of the method on samples from patients with two forms of gastrointestinal disease involving metabolic and microbial perturbation, CD and CoD [30, 31]. The developed method successfully detected and differentiated metabolic patterns of each group with a wide coverage. The method demonstrates strong cross-platform compatibility, with successful method application using two distinct analytical platforms, Orbitrap 240 LC-MS (ThermoFisher Scientific) and targeted triple-quadrupole (Shimadzu UK). This is valuable for future use of the method in laboratories using different technologies for metabolomic analysis.

While contradictive reports are found regarding metabolite extraction procedures, it is important to bring to light methods that are suitable in specific contexts to continue the drive towards standardisation. The use of biphasic extraction protocols is common in metabolomics sample preparation; however, method advancement must also reflect amenability to study design. A considerable amount of research [28, 32-34] suggests the importance of single-phase extraction procedures that can be used as a simpler, faster, and scalable alternatives to some of the more-extensive approaches, giving impetus for investigating the optimal monophasic extraction protocol for human faecal samples. Rapid and easy-to-use methods can greatly simplify metabolite extraction and thereby improve scalability and application in large clinical studies. In this sense, single-phase methods are advantageous as the single layer can easily be removed, minimizing the risk of sample loss and contamination. This is of paramount importance for large studies and those with limited sample amounts. Moreover, the method developed in this study uses fewer toxic chemicals and can therefore be deemed as more friendly to both the operator and environment.

In summary, untargeted and targeted LC-MS analysis of different extraction factors provides an insight into the specific methods which give the strongest metabolic output. Optimised sample pre-treatment and extraction methods ultimately improve protocol efficiency while simultaneously enhancing the MS signal obtained. Each small parameter change causes a small increase in the efficiency of LC-MS characteristics and so when combined, the accumulated difference in the overall protocol can result in a large improvement to the quality of data obtained. This is particularly important when using LCMS instrumentation of high sensitivity, and in doing so, the likelihood of matrix effects is also decreased. Furthermore, reproducibility of the method and the instrument are increased by documenting and working towards method standardisation. As the results from this study bring together some of the parameters of faecal metabolite extraction in agreement with existing studies, this supports evidence of an optimised and reproducible protocol that can be applied in a vast array of research and clinical settings. Moreover, the method covers a wide range of metabolites of different physiochemical properties to increase the capture of biological compounds. As an extension, employing the method to patients with gastrointestinal disease expands the protocol applicability to different sample types. This method addresses the requirement for affordable, reproducible, and environmentally friendly metabolite extraction protocols. Thus, the method described build on the foundations of protocol standardisation, allowing improved comparisons of future metabolomics studies using faecal samples.

## 5. Conclusions

Based on a series of optimisation experiments, we describe a protocol to extract metabolites from faecal samples for metabolomic analysis using an LC-MS system. We recommend the use of 50 mg freeze-dried faecal samples in a 1000 μL MeOH and bead beating extraction, as given by a reproducible increased metabolite measurement. The optimised faecal extraction method described here can be used for metabolomics investigations of a wide array of applications, with strong evidence for its suitability in studies of gastrointestinal disease. This contributes towards standardizing a framework of sample preparation, allowing easier and more accurate comparisons between studies.

## Data Availability

All data produced in the present study are available upon reasonable request to the authors.

## Author Contributions

“Conceptualisation, P.E.K, G.F, Z.R, K.G. and N.J.W.R.; methodology, P.E.K, H.J.N, S.M, Z.R, K.G and N.J.WR.; software, P.E.K, G.F and N.J.W.R.; validation, P.E.K, G.F and N.J.W.R.; formal analysis, .E.K, H.J.N, S.M, Z.R, K.G and N.J.WR; investigation, .E.K, H.J.N, S.M, and N.J.WR .; resources, R.K.R, R.H and K.G; data curation, P.E.K, G.F and N.J.W.R; writing—original draft preparation, .P.E.K and N.J.WR ; writing—review and editing, P.E.K, H.J.N, G.F, S.M, R.K.R, R.H, Z.R, K.G and N.J.W.R; visualisation, P.E.K and N.J.W.R; supervision, N.J.W.R, Z.R and K.G; project administration, N.J.W.R; funding acquisition, N.J.W.R, K.G and Z.R. All authors have read and agreed to the published version of the manuscript.”

## Institutional Review Board Statement

The study was conducted in accordance with the Declaration of Helsinki, and approved by the Institutional Review Board (or Ethics Committee) of NHS West of Scotland Research Ethics Committee (Ref: 11/WS/0006) for the study in patients with coeliac disease and the Yorkhill Research Ethics Committee (Reb: 05/S0708/66) for the study in patients with Crohn’s disease.

## Informed Consent Statement

Informed consent was obtained from all subjects involved in the study.

## Data Availability Statement

The data that support the findings of this study are available from the corresponding author upon reasonable request.

### Acknowledgments

N.J.W.R would like to acknowledge the Strathclyde Centre of Molecular Bio-science (www.scmb.strath.ac.uk) for access to LCMS instrumentation.

## References

Wishart, D.S.; Guo, A; Oler, E; Wang, F; Anjum, A; Peters, H; Dizon, R; Sayeeda, Z; Tian, S; Lee, B.L; Berjanskii, M; Mah, R; Yamamoto, M; Jovel, J; Torres-Calzada, C; Hiebert-Giesbrecht, M; Lui, V.W; Varshavi, D; Varshavi, D; Allen, D; Arndt, D; Khetarpal, N; Sivakumaran, A; Harford, K; Sanford, S; Yee, K; Cao, X; Budinski, Z; Liigand, J; Zhang, L; Zheng, J; Mandal, R; Karu, N; Dambrova, M; Schiöth, HB; Greiner, R; Gautam, V. HMDB 5.0: the Human Metabolome Database for 2022. Nucleic Acids Res. 2022. 50(D1): D622–D631. doi: 10.1093/nar/gkab1062.

Zierer, J; Jackson, M.A; Kastenmüller, G; Mangino, M; Long, T; Telenti, A; Mohney, R.P; Small, K.S; Bell, J.T; Steves, C.J; Valdes, A.M; Spector, T.D; Menni, C. The fecal metabolome as a functional readout of the gut microbiome. Nat Genet. 2018. 50(6):790–795. doi: 10.1038/s41588-018-0135-7.

Erben, V; Poschet, G; Schrotz-King, P; Brenner, H. Evaluation of different stool extraction methods for metabolomics measurements in human faecal samples. BMJ NPH. 2020. 4(2): 374–384. doi: 10.1136/bmjnph-2020-000202.

Coker, O. O; Liu, C; Wu, W. K. K; Wong, S. H; Jia, W; Sung, J. J. Y., & Yu, J. Altered gut metabolites and microbiota interactions are implicated in colorectal carcinogenesis and can be non-invasive diagnostic biomarkers. Microbiome. 2022. 10(1). doi: 10.1186/s40168-021-01208-5.

Yu, D; Du, J; Pu, X; Zheng, L; Chen, S; Wang, N; Li, J; Chen, S; Pan, S; Shen, B. The Gut Microbiome and Metabolites Are Altered and Interrelated in Patients With Rheumatoid Arthritis. Front Cell Infect Microbiol. 2022. 11: 763507. doi: 10.3389/fcimb.2021.763507.

Fang, Q; Liu, N; Zheng, B; Guo, F; Zeng, X; Huang, X; Ouyang, D. Roles of Gut Microbial Metabolites in Diabetic Kidney Disease. Front Endocrinol. 2021. 12: 636175. doi: 10.3389/fendo.2021.636175.

Zheng, L; Wen, X. L; Duan, S. L. Role of metabolites derived from gut microbiota in inflammatory bowel disease. World J. Clin. Cases. 2022. 10(9): 2660–2677. doi: 10.12998/wjcc.v10.i9.2660.

Di Cagno, R; Rizzello, CG; Gagliardi, F; Ricciuti, P; Ndagijimana, M; Francavilla, R; Guerzoni, ME; Crecchio, C; Gobbetti, M; De Angelis, M. Different fecal microbiotas and volatile organic compounds in treated and untreated children with celiac disease. Appl Environ Microbiol. 2009. 75(12): 3963–71. doi: 10.1128/AEM.02793-08.

Metwaly, A.; Dunkel, A.; Waldschmitt, N.; Raj, A. C. D.; Lagkouvardos, I.; Corraliza, A. M.; Mayorgas, A.; Martinez-Medina, M.; Reiter, S.; Schloter, M.; Hofmann, T.; Allez, M.; Panes, J.; Salas, A.; Haller, D. Integrated microbiota and metabolite profiles link Crohn’s disease to sulfur metabolism. Nat. Commun. 2020. 11(1): 4322. doi: 10.1038/s41467-020-17956-1.

Heinken, A.; Hertel, J.; Thiele, I. Metabolic modelling reveals broad changes in gut microbial metabolism in inflammatory bowel disease patients with dysbiosis. npj Syst Biol Appl. 2021. 7(1):19. doi: 10.1038/s41540-021-00178-6.

Khalkhal, E.; Rezaei-Tavirani, M.; Fathi, F.; Nobakht M Gh, BF.; Taherkhani, A.; Rostami-Nejad, M.; Asri, N.; Haidari, MH. Screening of Altered Metabolites and Metabolic Pathways in Celiac Disease Using NMR Spectroscopy. Biomed Res Int. 2021. 1798783. doi: 10.1155/2021/1798783.

Martín-Masot, R.; Mota-Martorell, N.; Jové, M.; Maldonado, J.; Pamplona, R.; Nestares, T. Alterations in one-carbon metabolism in celiac disease. Nutrients. 2020. 12(12), 1–14. doi: 10.3390/nu12123723.

Hosseinkhani, F.; Dubbelman, A. C.; Karu, N.; Harms, A. C.; Hankemeier, T. Towards standards for human fecal sample preparation in targeted and untargeted lc-hrms studies. Metabolites. 2021, 11(6). doi: 10.3390/metabo11060364.

Moosmang, S.; Pitscheider, M.; Sturm, S.; Seger, C.; Tilg, H.; Halabalaki, M.; Stuppner, H. Metabolomic analysis—Addressing NMR and LC-MS related problems in human feces sample preparation. Clinica Chimica Acta. 2019. 489, 169–176. doi: 10.1016/j.cca.2017.10.029.

Nandania, J.; Peddinti, G.; Pessia, A., Kokkonen, M.; Velagapudi, V. Validation and automation of a high-throughput multitargeted method for semiquantification of endogenous metabolites from different biological matrices using tandem mass spectrometry. Metabolites. 2018. 8(3). doi: 10.3390/metabo8030044.

Wu, J.; An, Y.; Yao, J.; Wang, Y.; Tang H. An optimised sample preparation method for NMR-based faecal metabonomic analysis. Analyst. 2010. 135: 1023–30. doi: 10.1039/b927543f.

Gholib, G; Heistermann, M; Agil, M; Supriatna, I; Purwantara, B; Nugraha, T.P; Engelhardt, A. Comparison of fecal preservation and extraction methods for steroid hormone metabolite analysis in wild crested macaques. Primates. 2018. 59(3):281–292. doi: 10.1007/s10329-018-0653-z.

Zhao, X.; Zhang, Z.; Hu, B.; Huang, W.; Yuan, C.; Zou, L. Response of gut microbiota to metabolite changes induced by endurance exercise. Front. Micro. 2018. 9:765. doi: 10.3389/fmicb.2018.00765.

de Zawadzki, A; Thiele, M; Suvitaival, T; Wretlind, A; Kim, M; Ali, M; Bjerre, A. F; Stahr, K; Mattila, I; Hansen, T; Krag, A; Legido-Quigley, C. High-Throughput UHPLC-MS to Screen Metabolites in Feces for Gut Metabolic Health. Metabolites. 2022. 12(3). doi: 10.3390/metabo12030211.

Want, E.J; O’Maille, G; Smith, C.A; Brandon, T.R; Uritboonthai, W; Qin, C; Trauger, S.A; Siuzdak, G. Solvent-dependent metabolite distribution, clustering, and protein extraction for serum profiling with mass spectrometry. Anal Chem. 2006. 78(3):743–52. doi: 10.1021/ac051312t.

Newton, J. M; Betts, E. L; Yiangou, L; Roldan, J. O; Tsaousis, A. D; Thompson, G. S. Establishing a metabolite extraction method to study the metabolome of blastocystis using nmr. Molecules. 2011. 26(11). doi: 10.3390/molecules26113285.

Deda, O; Gika, H. G; Wilson, I. D; Theodoridis, G. A. An overview of fecal sample preparation for global metabolic profiling. J. Pharm. Biomedical. 2015. 113: 137–150. doi: 10.1016/j.jpba.2015.02.006.

Zeng, M; Cao, H. Fast quantification of short chain fatty acids and ketone bodies by liquid chromatography-tandem massspectrometry after facile derivatization coupled with liquid-liquid extraction.J. Chromatogr. B Anal. Technol. Biomed. Life Sci. 2018. 1083, 137–14. doi: 10.1016/j.jchromb.2018.02.040.

Vuckovic, D. Current trends and challenges in sample preparation for global metabolomics using liquid chromatography-massspectrometry. Anal Bioanal Chem. 2012. 403, 1523–1548. doi: 10.1007/s00216-012-6039-y.

Fu, X; Calderón, C; Harm, T; Gawaz, M; Lämmerhofer, M. Advanced unified monophasic lipid extraction protocol with wide coverage on the polarity scale optimized for large-scale untargeted clinical lipidomics analysis of platelets. Analytica Chimica Acta. 2022. 1221, 340155. doi:10.1016/j.aca.2022.340155.

Cheng, K; Brunius, C; Fristedt, R; Landberg, R. An LC-QToF MS based method for untargeted metabolomics of human fecal samples. Metabolomics. 2020. 16(4). doi: 10.1007/s11306-020-01669-z.

Macioszek, S; Dudzik, D; Jacyna, J; Wozniak, A; Schöffski, P; Markuszewski, M. J. A robust method for sample preparation of gastrointestinal stromal tumour for LC/MS untargeted metabolomics. Metabolites. 2021. 11(8). doi: 10.3390/metabo11080554.

Southam, A. D; Pursell, H; Frigerio, G; Jankevics, A; Weber, R. J. M; Dunn, W. B. Characterization of Monophasic Solvent-Based Tissue Extractions for the Detection of Polar Metabolites and Lipids Applying Ultrahigh-Performance Liquid Chromatography-Mass Spectrometry Clinical Metabolic Phenotyping Assays. J. Proteome Res. 2021. 20(1), 831–840. doi: 10.1021/acs.jproteome.0c00660.

Sumner, L.W; Amberg. A; Barrett, D; Beale, M.H; Beger, R; Daykin, C.A; Fan, T.W; Fiehn, O; Goodacre, R; Griffin, J.L; Hankemeier, T; Hardy, N; Harnly, J; Higashi, R; Kopka, J; Lane, A.N; Lindon, J.C; Marriott, P; Nicholls, A.W; Reily, M.D; Thaden, J.J; Viant, M.R. Proposed minimum reporting standards for chemical analysis Chemical Analysis Working Group (CAWG) Metabolomics Standards Initiative (MSI). Metabolomics. 2007. 3(3):211–221. doi: 10.1007/s11306-007-0082-2.

Zafeiropoulou, K; Nichols, B; Mackinder, M; Biskou, O; Rizou, E; Karanikolou, A; Clark, C; Buchanan, E; Cardigan, T; Duncan, H; Wands, D; Russell, J; Hansen, R; Russell, R.K; McGrogan, P; Edwards, C.A; Ijaz, U.Z; Gerasimidis K. Alterations in Intestinal Microbiota of Children With Celiac Disease at the Time of Diagnosis and on a Gluten-free Diet. Gastroenterology. 2020. 159(6):2039-2051.e20. doi: 10.1053/j.gastro.2020.08.007.

Quince, C; Ijaz, U.Z; Loman, N; Eren, A.M; Saulnier, D; Russell, J; Haig, S.J; Calus, S.T; Quick. J; Barclay, A; Bertz, M; Blaut, M; Hansen, R; McGrogan, P; Russell, R.K; Edwards, C.A; Gerasimidis K. Extensive Modulation of the Fecal Metagenome in Children With Crohn’s Disease During Exclusive Enteral Nutrition. Am J Gastroenterol. 2015. 110(12):1718–29. doi: 10.1038/ajg.2015.357.

Medina, J; van der Velpen, V; Teav, T; Guitton, Y; Gallart-Ayala, H; Ivanisevic, J. Single-step extraction coupled with targeted hilic-ms/ms approach for comprehensive analysis of human plasma lipidome and polar metabolome. Metabolites. 2020. 10(12), 1–17. doi: 10.3390/metabo10120495.

Alshehry, Z. H; Barlow, C. K; Weir, J. M; Zhou, Y; McConville, M. J; Meikle, P. J. An efficient single phase method for the extraction of plasma lipids. Metabolites. 2015. 5(2), 389–403. doi: 10.3390/metabo5020389.

Peterson, A.L; Walker, A.K; Sloan E.K; Creek D.J. Optimized Method for Untargeted Metabolomics Analysis of MDA-MB-231 Breast Cancer Cells. Metabolites. 2016. 6(4):30. doi: 10.3390/metabo6040030.

